# Development of Novel Measures for Alzheimer’s Disease Prevention Trials (NoMAD)

**DOI:** 10.1101/2021.01.13.21249627

**Authors:** Sophie A. Bell, Hannah R. Cohen, Seonjoo Lee, Hyun Kim, Adam Ciarleglio, Howard Andrews, Andres M. Rivera, Kay Igwe, Adam M. Brickman, D.P. Devanand, Philip D. Harvey, Lon S. Schneider, Terry E. Goldberg

## Abstract

**Introduction:** Assessment of cognition and everyday function is essential in clinical trials for Alzheimer’s disease (AD). Two novel measures of cognition (No Practice Effects (NPE) cognitive battery and Miami Computerized Functional Assessment Scale (CFAS)) were designed to have robust psychometric properties and reduced practice and ceiling effects. This study aims to evaluate if the NPE and CFAS demonstrate stronger psychometric properties and reduced practice effects compared with established measures, including the Preclinical Alzheimer Cognitive Composite (PACC), Alzheimer’s Disease Assessment Scale-Cognitive Subscale (ADAS-Cog), and Functional Activities Questionnaire (FAQ).

**Methods:** This parallel group, four-site study will randomize 320 cognitively intact adults aged 60 to 85 years to novel or well-established measures of cognition and function. All participants will receive assessments at baseline (week 0), week 12, and week 52, as well as a brain MRI scan and Apolipoprotein E genetic test at study entry. Analyses will determine psychometric properties of the NPE and CFAS, compare the sensitivity of measures to AD risk markers, and identify cognitive domains within the NPE.

**Discussion:** Practice effects have been a major limitation of Alzheimer’s disease clinical trials that typically assess cognitive changes over serial assessments. Detection of functional impairment in cognitively normal individuals with biomarkers for Alzheimer’s disease requires instruments sensitive to very subtle functional changes. This study is intended to support the validation of two new composite measures, the NPE battery and the CFAS, which may advance clinical testing of interventions for individuals across the spectrum of early stage Alzheimer’s disease.

**Trial Registration:** NCT03900273

## INTRODUCTION

Accurately detecting cognitive impairment/decline and everyday functioning in preclinical and early mild cognitive impairment (MCI) adults is critical in clinical trials assessing potential treatment of Alzheimer’s disease (AD). Clinical trials often use serial testing designs that assess cognition on several occasions within a limited period. In this structure, cognitive performance in randomized placebo or active treatment groups are compared in terms of the rate of change. This serial testing may result in reduced sensitivity to treatment by the induction of practice effects. Practice-related improvements in serial testing interfere with the detection of cognitive enhancement or subtle decline because they reduce differences between treatment groups, do not generalize or transfer readily, and are item- or paradigm specific. The MELODEM initiative for methodological advances in AD longitudinal studies and clinical trials pinpointed practice effects as a major concern.^1^ Subtle changes in everyday functioning are also difficult to measure with most established measures. Global informant report rating measures commonly used to characterize functional decline in late MCI and clinical AD are unlikely to detect very subtle functional changes in cognitively normal persons with amyloid or other biomarkers. Therefore, a sensitive set of tests assessing important cognitive and functional domains, with good psychometric properties, and resistant to practice effects would accurately monitor cognitive function and subtle declines or improvements over time.

Two novel measures of cognition and everyday functioning, the No Practice Effect (NPE) Battery and the Miami Computerized Functional Assessment Scale (CFAS)^2^ were developed to overcome the limiting features of prior instruments used in clinical trials for preclinical AD. The NPE battery was constructed using principles from the cognitive science literature that potentially substantially reduce practice- and ceiling effects (e.g., alternative forms, distractors to reduce memorization of responses). In the CFAS, computer-delivered simulations assess cognitively complex functional skills required for independent living and sensitive to early decline.

The present study aims to examine psychometric characteristics of two novel measures (NPE and CFAS), in comparison to a set of established measures (Preclinical Alzheimer Cognitive Composite (PACC),^3^ the Alzheimer’s Disease Assessment Scale–Cognitive Subscale (ADAS-Cog),^4^ and the Functional Activities Questionnaire (FAQ)).^5^ We hypothesize that these novel measures will demonstrate reduced practice effects compared to established measures while demonstrating adequate test retest reliabilities, coefficient of variation (CV), equivalence of alternate forms, and minimal ceiling or floor effects. The second aim of the study is to compare the sensitivity of the novel and established measures by contrasting performances by subgroups defined by AD biomarkers and genetic factors. We hypothesize that the NPE and CFAS will demonstrate larger differences between biomarker derived subgroups than the established measures. As an exploratory measure, we will determine whether practice effects - assessed as slope from baseline to endpoint - are related to hippocampal volume within the established measures group. The exploratory aim of the study is to examine what different cognitive domains within the NPE (e.g., episodic memory, working memory/ speed, executive function, attentional monitoring) will have differential relationships with biomarkers, risk markers, and CFAS functional tasks.

## METHODS AND ANALYSIS

### Study Design

This study uses a novel randomized, parallel group design to compare neuropsychological test batteries. One group receives novel measures and the other receives established measures. The study takes place over the course of 12 months. In-clinic visits take place at baseline (week 0), week 12, and week 52. The same assessment battery is administered throughout these timepoints depending on random assignments to one of the two groups. Thus, novel measures are validated within a clinical trials armature in which participants are randomly assigned to a novel measures or established measures group that then undergoes serial assessment. In effect, this is a novel “intent-to-test” design. This study design reflects the structure and methods of a clinical trial and allows assessment of differences in outcomes as a function of test battery. It eliminates potential interference effects between established and novel measures, especially those involving verbal memory. Additionally, by assessing several risk markers for neurodegeneration, including hippocampal volume, cortical thickness, along with APOE genotype, this study will assess the relative associations of novel and established measures to these established biomarkers. This project is registered on ClinicalTrials.gov as Development of Novel Measures for Alzheimer’s Disease Prevention Trials (NoMAD); ClinicalTrials.gov Identifier: NCT03900273.

### Study Participants: Recruitment and Eligibility

For the current study, we aim to enroll 320 cognitively intact older adults ranging in age from 60 to 85. The participants are recruited across four sites including the New York State Psychiatric Institute/ Columbia University Irving Medical Center (NYSPI), Litwin-Zucker Alzheimer’s Research Center/Feinstein Institute for Medical Research, University of Miami – Miller School of Medicine, and University of Southern California – Keck School of Medicine. Each site is expected to recruit 80 participants.

Detailed inclusion/exclusion criteria are listed in Table 1. Because the current study aims to enroll non-cognitively impaired older adults, individuals are screened for potential impairment based on two tests measuring general cognitive ability, the Folstein Mini-Mental State Examination (MMSE)^6^ and Wechsler Memory Scale-III Logical Memory Story A (Logical Memory).^7^ Individuals are also screened for history of various psychiatric, neurologic, and other medical conditions that could impact cognition.

**Table 1.** Inclusion/Exclusion Criteria

| Inclusion Criteria | Exclusion Criteria |
| --- | --- |
| 1. English speaking, 60-85 years of age (inclusive). | 1. Diagnosis of stroke or excessive risk of cerebrovascular disease (CVD), as defined by a score of 4 or more on the Hachinski Ischemic Scale |
| 2. Mini Mental State Examination<sup>6</sup> $\geq 24$ | 2. Neurologic disease including movement disorders, multiple sclerosis, epilepsy, and TBI (with greater than 15 minutes loss of consciousness). |
| 3. Wechsler Memory Scale-III Logical Memory<sup>7</sup> delayed recall score $\geq 9$ ($\geq 5$ for participants with 8-15 years of education; $\geq 3$ for participants with 0-7 years of education) | 3. Untreated diabetes |
| 4. A family member or other individual who is in contact with the participant and consents to serve as informant during the study (can be a telephone informant for participants who do not have a live-in informant) | 4. Current DSM-5 Axis I psychiatric diagnosis of schizophrenia, schizoaffective disorder, or bipolar disorder; current major depression as determined by a Geriatric Depression Scale $> 5$; current alcohol or substance use disorder |
|  | 5. Active treatment of cancer |
|  | 6. Women who are pre-menopausal and are pregnant |
|  | 7. Use of antidepressants with large anticholinergic properties, including amitriptyline, amoxapine, clomipramine, desipramine, doxepin, imipramine, isocarboxazid, lithium, maprotiline, mirtazapine, nortriptyline, tranylcypromine trimipramine, and phenelzine. |

Randomization of participants to either the novel measures or well-established measures is implemented before the baseline visit. This is a non-blinded study. Within each site, participants are stratified by age group (60-72, 73-85) and randomly assigned to test type in a 1:1 allocation, based on a pseudorandom algorithm developed and housed at NYSPI.

### Study Measures

Study measures are listed in Table 2 with the time points at which they are administered. All in-person measures will be administered by trained research coordinators. The 15-item Geriatric Depression Scale (GDS)^8^ will be used to assess depressive symptoms at the screening visit, 3-month visit, and 12-month visit for both the well-established and novel measures groups. If the GDS is greater than 5 at any visit, the patient will be evaluated by a psychiatrist and an appropriate clinical referral will be made for treatment of depression.

**Table 2.** Study Procedures

| Measure | Arm 1 (Well-Established Measures) |  |  |  | Arm 2 (Novel Measures) |  |  |  |
| --- | --- | --- | --- | --- | --- | --- | --- | --- |
|  | Screen | Baseline | 3-month | 12-month | Screen | Baseline | 3-month | 12-month |
| Demographics History | X |  |  |  | X |  |  |  |
| Inclusion/Exclusion Form | X |  |  |  | X |  |  |  |
| Informed Consent | X |  |  |  | X |  |  |  |
| Concomitant Medications | X | X | X | X | X | X | X | X |
| Vitals |  | X | X | X |  | X | X | X |
| ApoE and Blood Test | X |  |  |  | X |  |  |  |
| MRI Scan of Brain | X |  |  |  | X |  |  |  |
| Geriatric Depression Scale<sup>8</sup> | X |  | X | X | X |  | X | X |
| MMSE<sup>6</sup> | X |  | X | X | X |  |  |  |
| Logical Memory<sup>7</sup> | X |  | X | X | X |  |  |  |
| Selective Reminding Test<sup>9</sup> |  | X | X | X |  |  |  |  |
| Digit Symbol Coding<sup>10</sup> |  | X | X | X |  |  |  |  |
| ADAS-Cog<sup>4</sup> |  | X | X | X |  |  |  |  |
| FAQ<sup>5</sup> |  | X | X | X |  |  |  |  |
| N-back test<sup>11</sup> |  |  |  |  |  | X | X | X |
| Simple Letter-Number Span<sup>12</sup> |  |  |  |  |  | X | X | X |
| Executive Letter-Number Span<sup>12</sup> |  |  |  |  |  | X | X | X |
| Word RMT<sup>17</sup> |  |  |  |  |  | X | X | X |
| Symbol Coding<sup>15</sup> |  |  |  |  |  | X | X | X |
| Verbal Fluency<sup>16</sup> |  |  |  |  |  | X | X | X |
| CFAS<sup>2</sup> |  |  |  |  |  | X | X | X |
| Brown-Peterson<sup>13 14</sup> |  |  |  |  |  | X | X | X |

MMSE<sup>6</sup>, Mini-Mental State Exam; Logical Memory<sup>7</sup>, Wechsler Memory Scale-III Logical Memory Story A; ADAS-Cog<sup>4</sup>, Alzheimer's Disease Assessment Scale – Cognitive Subscale 11; FAQ<sup>5</sup>, Functional Activities Questionnaire; Word RMT<sup>17</sup>, Word Recognition Memory Test; CFAS<sup>2</sup>, University of Miami Computerized Functional Assessment Scale.

#### Well-Established Measures

The PACC^3^ composite score includes the Logical Memory Delayed Recall,^7^ Total Recall from Selective Reminding,^9^ Digit Symbol Coding,^10^ and MMSE.^6^ The MMSE and Logical Memory will not be administered at baseline as participant scores from the screening visit will be used for baseline. These two measures will subsequently be administered at 3-month and 12-month visits for those in the well-established group. Additionally, we will employ the ADAS-Cog,^4^ a widely used measure of cognition in clinical trials of AD, MCI, and prodromal AD. The 11-item ADAS-Cog will be administered to participants at each time point. Lastly, the FAQ,^5^ an informant-based measure of everyday function, will be conducted either in-person or by telephone at each time point as well.

#### Novel Measures

For the novel test-battery group, participants receive the NPE and the CFAS.^2^ As there are alternate forms for the NPE and the CFAS, those in the novel measures group also receive a randomization sequence determining which form is administered at each time point.

The order of the three alternate forms of the NPE and CFAS are counterbalanced across subjects in this group (e.g., Form A then B then C to subject 1; form B then A then C to subject 2, etc.). Once the tests are completed and scored, two composite scores will be derived from each of the NPE battery and the CFAS.

#### No Practice Effect (NPE) Test Battery

The NPE battery was constructed using principles that potentially substantially reduce practice- and ceiling effects. Tests of working memory, attention, and executive function were designed with multiple items, a restricted set of stimuli that reduced the chance of frank memorization of responses between sessions, and alternative and equivalent forms with different items and sequences in tests. For episodic memory, obligatory common encoding of items was included to reduce strategy changes, followed by testing of recognition and the use of alternate forms. The majority of the NPE subtests are computerized or partially computerized. All tests have three equivalent alternate forms. The NPE subtests include the N-Back (Goldberg et al. 2003),^11^ Simple Letter Number Span, Executive Letter Number Span,^12^ Brown-Peterson,^13 14^ Symbol Coding,^15^ Verbal Fluency,^16^ and Word Recognition Memory Test (Word RMT).^17^ Detailed subtest descriptions are presented in Table 3 along with associated cognitive domains. The two conditions of the N-Back test are displayed in Figure 2.

**Table 3:**
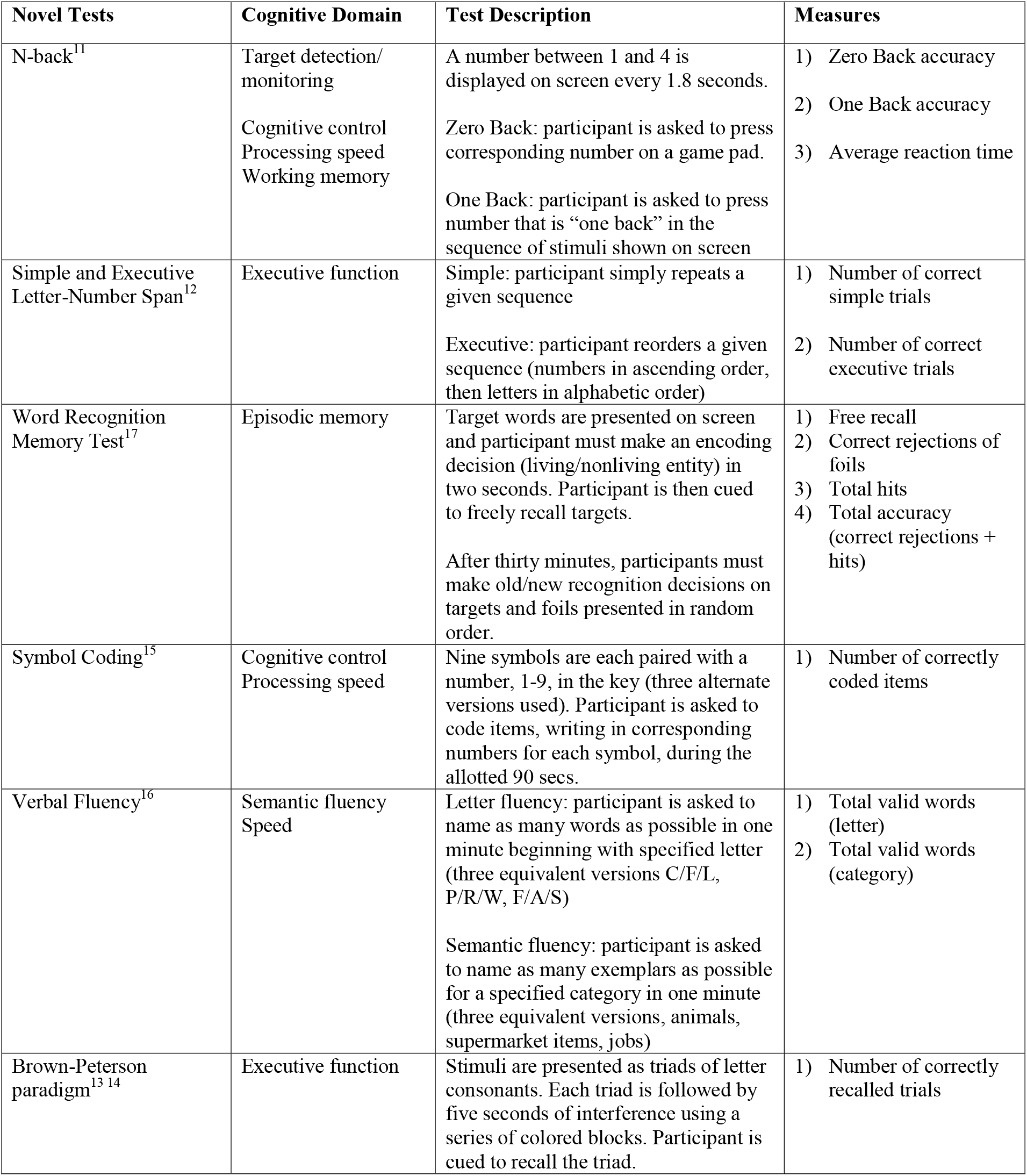
Cognitive Domains and Descriptions of No Practice Effect (NPE) test battery

| **Novel Tests** | **Cognitive Domain** | **Test Description** | **Measures** |
| --- | --- | --- | --- |
| N-back<sup>11</sup> | Target detection/<br>monitoring<br>Cognitive control<br>Processing speed<br>Working memory | A number between 1 and 4 is displayed on screen every 1.8 seconds.<br>Zero Back: participant is asked to press corresponding number on a game pad.<br>One Back: participant is asked to press number that is “one back” in the sequence of stimuli shown on screen | 1) Zero Back accuracy<br>2) One Back accuracy<br>3) Average reaction time |
| Simple and Executive Letter-Number Span<sup>12</sup> | Executive function | Simple: participant simply repeats a given sequence<br>Executive: participant reorders a given sequence (numbers in ascending order, then letters in alphabetic order) | 1) Number of correct simple trials<br>2) Number of correct executive trials |
| Word Recognition Memory Test<sup>17</sup> | Episodic memory | Target words are presented on screen and participant must make an encoding decision (living/nonliving entity) in two seconds. Participant is then cued to freely recall targets.<br>After thirty minutes, participants must make old/new recognition decisions on targets and foils presented in random order. | 1) Free recall<br>2) Correct rejections of foils<br>3) Total hits<br>4) Total accuracy (correct rejections + hits) |
| Symbol Coding<sup>15</sup> | Cognitive control<br>Processing speed | Nine symbols are each paired with a number, 1-9, in the key (three alternate versions used). Participant is asked to code items, writing in corresponding numbers for each symbol, during the allotted 90 secs. | 1) Number of correctly coded items |
| Verbal Fluency<sup>16</sup> | Semantic fluency<br>Speed | Letter fluency: participant is asked to name as many words as possible in one minute beginning with specified letter (three equivalent versions C/F/L, P/R/W, F/A/S)<br>Semantic fluency: participant is asked to name as many exemplars as possible for a specified category in one minute (three equivalent versions, animals, supermarket items, jobs) | 1) Total valid words (letter)<br>2) Total valid words (category) |
| Brown-Peterson paradigm<sup>13 14</sup> | Executive function | Stimuli are presented as triads of letter consonants. Each triad is followed by five seconds of interference using a series of colored blocks. Participant is cued to recall the triad. | 1) Number of correctly recalled trials |

**Figure 1.**
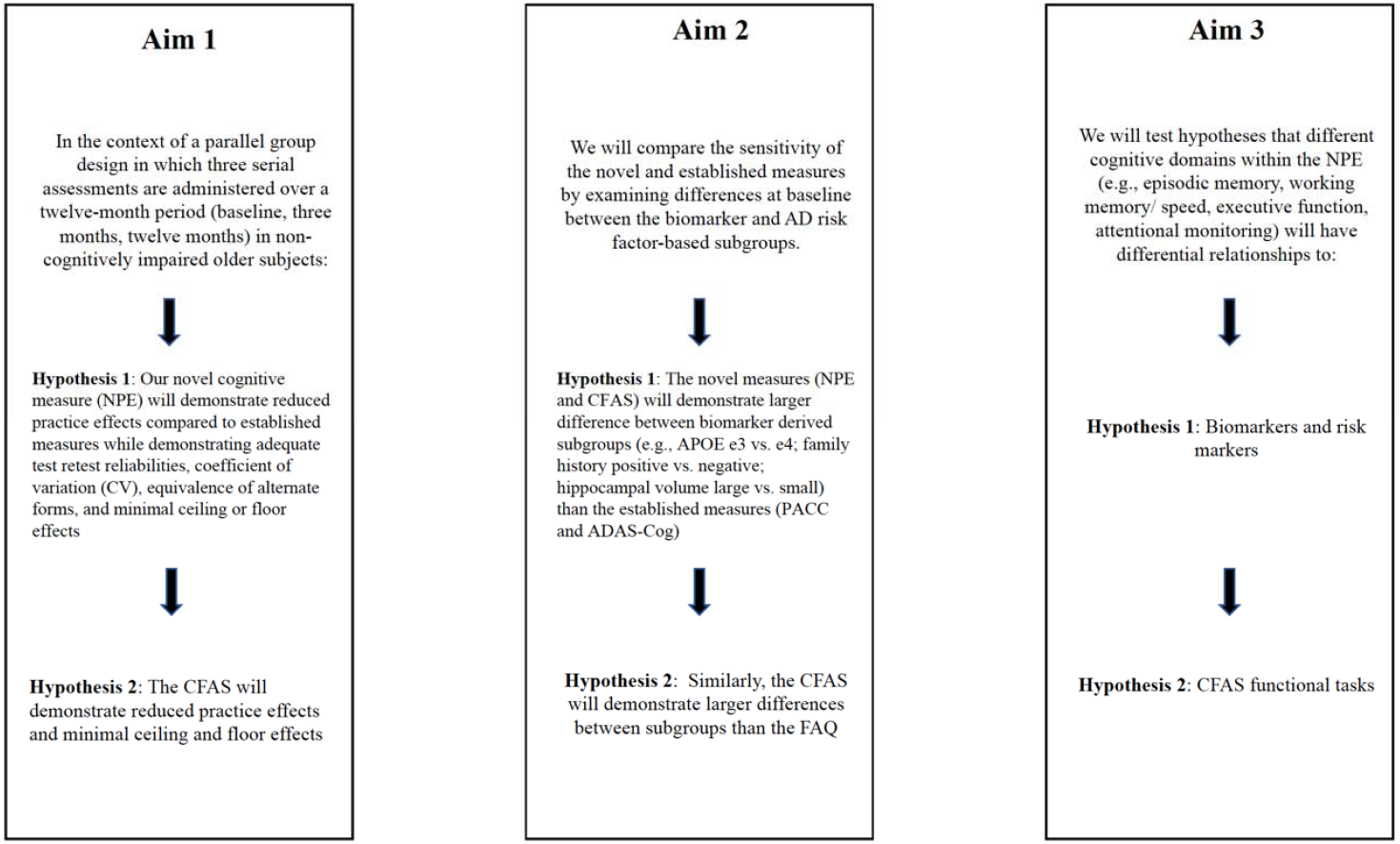
Aims and Hypotheses

**Figure 2.**
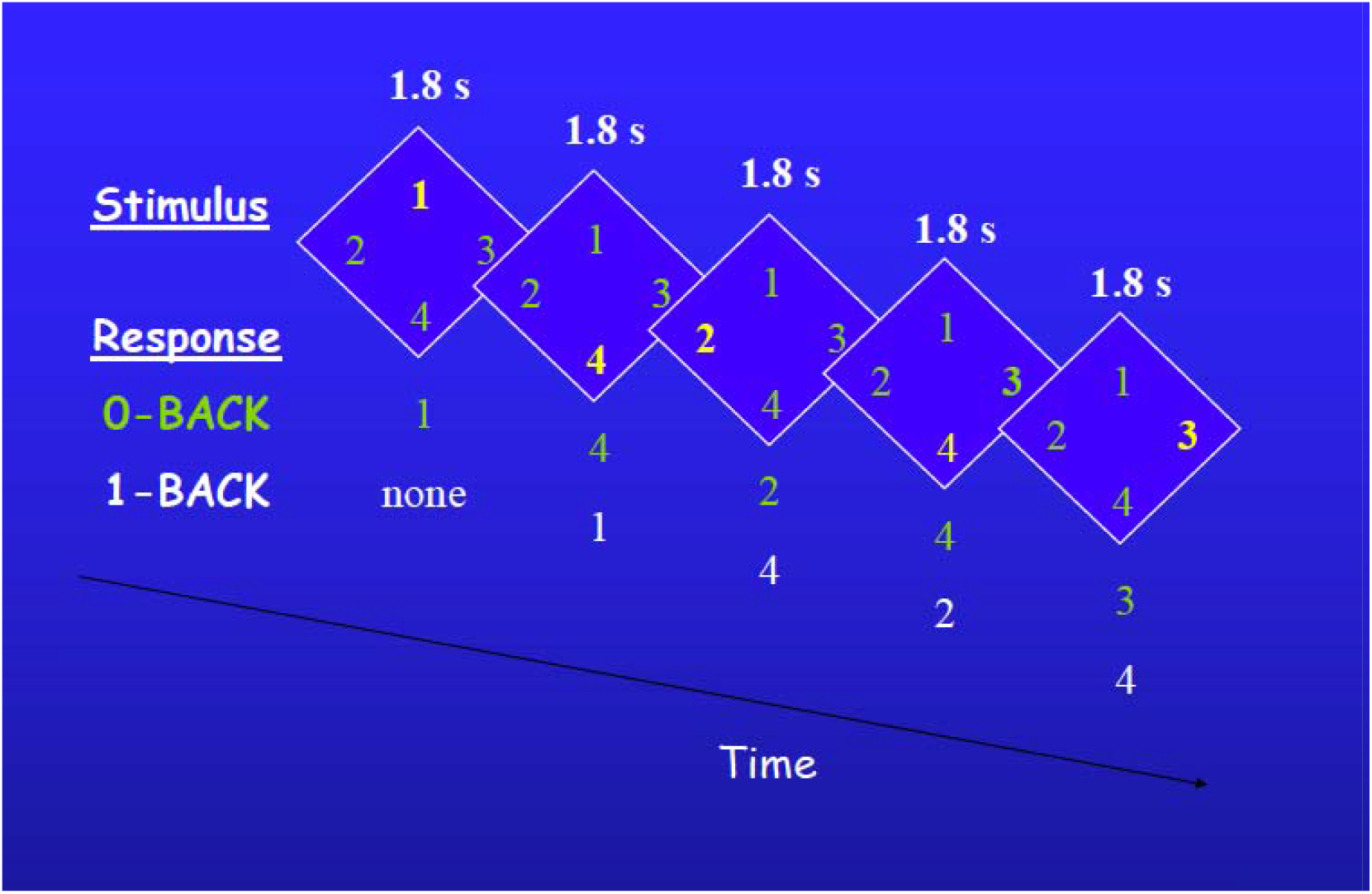
N-Back Task

#### Miami Computerized Functional Assessment Scale (CFAS)^2^

The CFAS is a set of 4 computer-delivered simulations (Figure 3). The simulations are realistic analogues of actual functional skills required for independent living and have alternative forms that maintain the same general task demands. The first is a virtual ATM banking session during which participants enter their PIN, check their balance, transfer money, and make a withdrawal, among other tasks. The second simulates an online banking session. The ticket kiosk simulation involves purchasing single ride and longer-term tickets, as well as checking schedules and adding money to a 2-week tourist metro-card. The final module, medication management, simulates organizing medications into pill boxes over the course of a week and answering comprehension questions based on medication labels. In addition to accuracy variables, these measures also include completion time variables that could capture subtle changes in processing speed.

**Figure 3.**
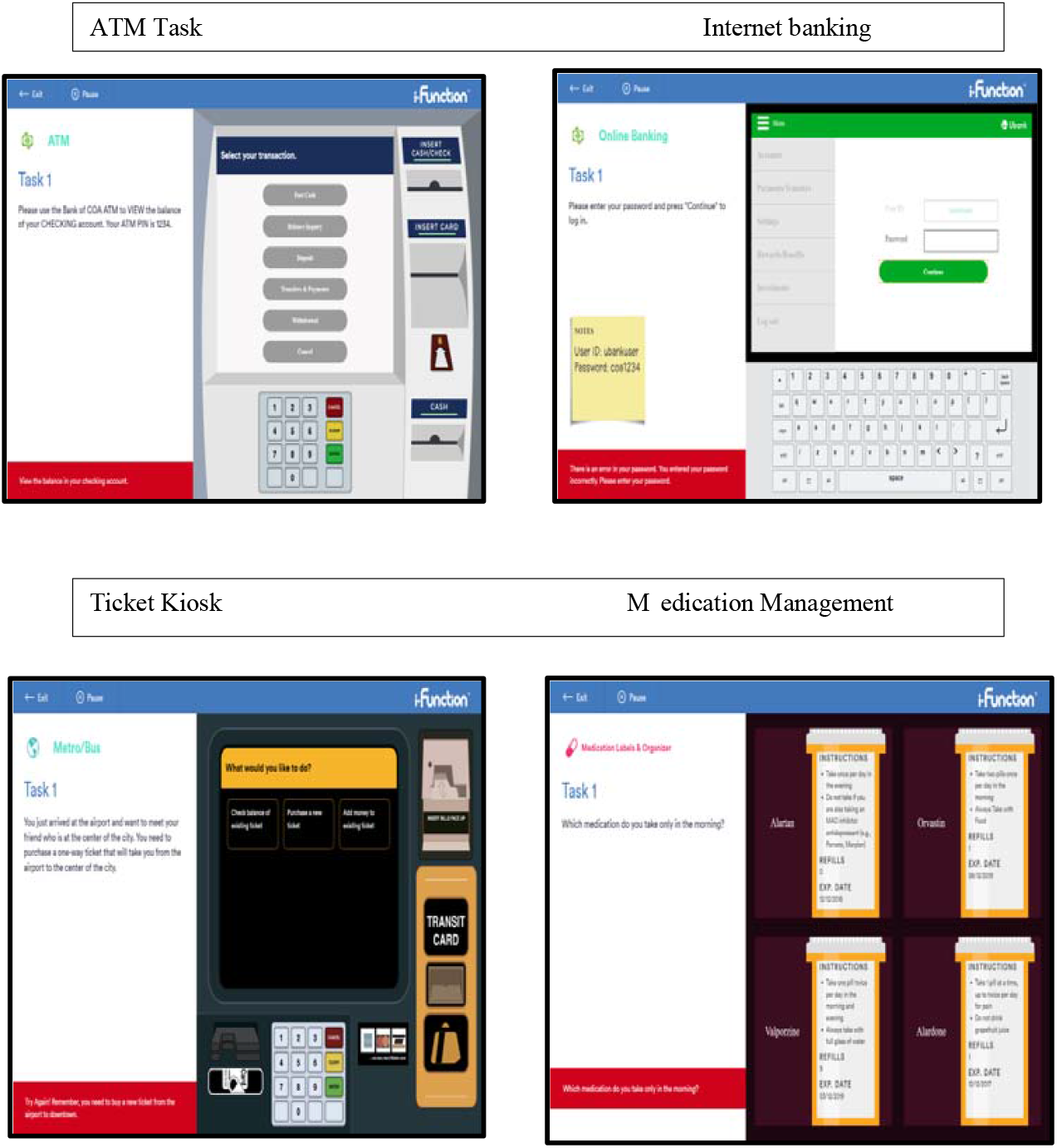
Miami Computerized Functional Assessment Scale

### Genetic Risk Marker

Apolipoprotein E (ApoE) genetic analysis will be done via DNA extraction on a blood sample through the laboratory of the Human Genetics Resources Core at Columbia University Medical Center.

### Structural MRI

High-resolution T1-weighted magnetic resonance imaging (MRI) will be acquired at each site in order to quantify regional volume and cortical thickness. Using each individual’s T1-weighted image, structural imaging measures of both global and regional brain volume and regional measures of cortical thickness are derived with FreeSurfer v6.0 http://surfer.nmr.mgh.harvard.edu/). Bilateral hippocampal volume corrected for intracranial volume will be the primary volumetric measure. Cortical thickness will also be assessed as specific patterns of cortical thinning are found in AD.^18 19^

### Data Management

The Columbia University Data Coordinating Center has designed and implemented a data management system for this study using REDCap technology.^20 21^ Data is transcribed onto paper forms for each participant at each visit. Authorized staff at each site including program managers, research coordinators, and data personnel are then able to access the online REDCap database and enter all data. Quality control mechanisms include automated checks (ensuring that each entered value is within the pre-specified range for each field) and manual data entry error checks by study personnel. Forms from the study database are then downloaded and provided to project statisticians as statistical system files (e.g., SPSS and SAS). Each study site maintains the link between participant contact information and the study ID used in the database and can only see, enter, and edit data from that site. To ensure confidentiality, the online data system does not contain explicit participant identifiers.

### Patient and Public Involvement

Patients and the public were not involved in the design and conduct of the study.

### Statistical Approaches

#### Statistical Analysis

Before the specific statistical techniques are applied, all variables will be examined at all time points for outliers and inconsistencies. Additionally, if there are significant differences between randomized groups with respect to site, sex, or education, these variables will be adjusted in the primary analyses. When assessing the relationships between MRI measures and test outcomes, site and magnet type will be adjusted. All raw test scores will be converted to z-scores referenced to baseline test scores.

To examine the test effect (Aim 1), we will use a linear mixed effects model with composite score of the cognitive domains as the dependent variable; test, time and their interactions as the fixed effects; and a random intercept to account for with-in subject correlation due to repeated measurement. All models will be adjusted for sex, age, education, or site. If the F-test for time x test interaction is significant, we will conduct post-hoc contrast analysis. For the novel tests, we will compute the Cohen’s d (and 95% confidence intervals (CIs)) between baseline and 12-month measures in order to determine if they are smaller than 0.15. We will examine test-retest reliability between baseline and 3-month measures by computing Pearson correlations and corresponding 95% CIs. Reliabilities greater than 0.70 will be considered adequate. Measures of dispersion, including skewness, kurtosis, and ceiling and floor values will also be examined. The proportion of participants at ceiling for each test will be calculated, and Chi-squared tests will be used to test for differences in those proportions between the novel and established tests. One-way ANOVA will be used to test for differences in un-standardized scores on alternate forms of the novel tests which were designed to be equivalent.

To assess differences in scores between biomarker-based and AD risk factor-based groups (Aim 2), we will use three separate multiple linear regression models with baseline test score as the dependent variable, and with test, each of the biomarker/risk-factor subgroup indicators (one in each model), and their interaction terms as the dependent variables.

We will conduct a confirmatory factor analysis to identify and describe different cognitive domains within the NPE (e.g., episodic memory, working memory/ speed, executive function cognitive control, attentional monitoring, fluency). Measures for each domain for the primary confirmatory factor analysis are described in Table 3. The factor loadings will be computed using the full information maximum likelihood method, and the factor scores will be computed. We will test the coefficients for each primary predictor to assess association with the specified domain score. For a given domain score, we will fit separate models for each marker as well as a combined model containing all markers. Similarly, we will consider CFAS functional measures as primary predictors for NPE domain scores and an NPE composite score.

#### Sample Size

The sample size is 320 (160 in each arm), and 3 measurements (0, 3, 12 months) are obtained for each participant. We conservatively assume that 10% of the sample will drop out by month 12. We conducted a power analysis using the RMASS program for longitudinal studies; the smallest effect size at 12 months between the two test groups can be detected with 80% power at an overall 5% significance level. Assuming the correlation between repeated measures is r=0.3 (moderate correlation), and with the variance of the random intercept set to be 1, the smallest detectable effect size is d=0.26 Hence, we are adequately powered to detect differences in the two test groups. Next, with 320 participants we have >80% power to detect the test by subgroup interaction with Cohen’s f^2^> 0.03 (small effect size)^22^ using two-sided 0.05-level significance tests. Lastly, with 160 participants in the novel test group, we have >80% power to detect associations with Cohen’s f^2^> 0.05 (small effect size) between biomarker/risk marker measures and a given NPE domain score using two sided 0.05-level significance tests.

### Ethics and Dissemination

This study protocol has been approved by the Institutional Review Board (IRB) at each site. All participants are required to provide written informed consent. During the informed consent process, participants are told that the information they provide and their test outcomes will be kept strictly confidential. All participant data are kept securely in the REDCap database system. Only research personnel with specific permissions have access to REDCap and any identifying information, except in some cases when audits are performed by either State or Federal regulatory personnel.

Participants are informed of possible albeit unlikely side effects of cognitive testing or blood draw. Local bruising and discomfort are the most common side effects associated with venipuncture. Participants are permitted to take breaks if fatigued from cognitive testing at any point. Risks of being involved in genetic testing include the misuse of personal, genetic information. All personnel who will have access to genetic information about the participants are ethically and legally obligated to maintain the confidence of that information. Genetic information is not being used to enroll participants nor will it be disclosed to participants. The MRI procedure is also considered no greater than minimal risk because we are not using contrast agents or an experimental high-field strength magnet. Overall, the knowledge to be gained from this study is substantial with little potential risks detracting from benefits. While there is no direct benefit to participants, the study may have salutary and broad consequences for clinical trials in the field of AD.

With regard to dissemination, study findings will be shared with the academic community and the community at large through peer-reviewed publications, conferences, and public websites, including clinicaltrials.gov.

## DISCUSSION

There is an unmet need for studies to validate trial outcome measures with updated norms and complete psychometric information. The sensitivity and difficulty levels of assessments used in clinical trials are two critical factors that need to be addressed to quantify cognitive changes across different phases of AD.

Practice effects have been a major limitation of AD longitudinal studies and clinical trials that typically assess cognitive changes over multiple serial assessments. Practice effects can mask treatment effects by increasing variance in endpoints. Some individuals will demonstrate greater practice effects than others, and statistically, this will yield a reduction in between-group effect size by increasing the pooled standard deviation and potentially reducing the mean group differences in outcome measures. As a result, statistical power will be reduced.

It is now clearly established that healthy older adults can generate significant practice effects (Cohens d = 0.25) over two to three assessments.^23^ Notably, Matthews and colleagues^24^ used the National Alzheimer’s Coordinating Center (NACC) Uniform Data Set^25^ cognitive battery and compared performance of cognitively normal, MCI, and dementia subgroups with prior test exposure to those who were naiLve to testing. They found that scores of test-exposed participants were greater than scores of test-naïve participants, in both cognitively normal and MCI groups. Strikingly, they found significant practice effects (Cohen’s d nearly.40) in both the older healthy control group and the MCI group on the composite cognitive measure that included multiple cognitive domains spanning attention, executive function, memory, and language. These data support the contention that practice effects are prevalent in even cognitively impaired individuals. Further, ceiling effects are especially prominent in healthy populations on cognitive measures such as orientation. Orientation is often tested in mental status exams, including the MMSE.^6^ Schneider and Goldberg^26^ found that approximately 80% of participants score at ceiling and another 17% score 9/10 on MMSE orientation items in the Alzheimer’s Disease Neuroimaging Initiative (ADNI) data set. Interestingly, approximately 42% of subjects with amnestic MCI also scored at ceiling.

In healthy or pre-clinical populations, established measures commonly used to detect functional impairment in late MCI and clinical AD are unlikely to capture subtle changes in everyday functioning. For instance, the FAQ^5^, an informant-based measure of everyday function, focuses on instrumental activities of daily living (ADLs) in mild-moderate dementia. In a large ADNI database, the majority of cognitively normal older adults obtained a score of zero on the FAQ, indicating a floor effect.^27^ Another informant-based functional measure, namely the Alzheimer’s Disease Cooperative Study–Activities of Daily Living Inventory (ADCS-ADL), also demonstrated skewed distribution and high ceiling effect (28% in controls).^28^ Because ADL’s are generally preserved in pre-clinical stages of AD, application of informant-based functional measures to asymptomatic individuals may not accurately detect subtle cognitive changes that may be present with AD biomarkers. While a performance-based measure of everyday functioning, the UCSD Performance-based Skills Assessment (UPSA)^29^ has a lower ceiling effect and reveals a greater contrast between cognitively healthy participants and those with MCI, these measures are still vulnerable to practice effects.^30^

Computerized functional tasks are needed because paper and pencil functional capacity measures, including the Financial Capacity Instrument-Short Form (FCI-SF)^31^ and the UPSA, have subtests that require performance of everyday tasks that may be becoming outdated (e.g., dialing directory assistance, writing paper checks, and making paper check deposits). The novel tasks have demonstrated sensitivity to age-related differences in healthy adults, strong correlations with cognitive test performance, and distinction of the performance of healthy people from various cognitively impaired populations, including amnestic MCI.^2 32^ Further, the computer-delivered functional tasks avoid biases often associated with informant-based and self-report measures.^2 32^

### Why use composite scales?

Two composite scores will be derived from the NPE battery and the CFAS. Composite tests combine several clinical subtests, often covering multiple domains, and then derive a single outcome score from their averages. Composite scores can be effective in monitoring outcomes in clinical trials in that they provide for an aggregate score that reflects the general cognitive architecture.^26^ Compared to individual tests, composites may also improve test-retest reliability by involving a larger item pool. However, few cognitive and functional composites thus far have sufficient psychometric data to suggest that they are truly an improvement on individual scales.

A recent review paper by Schneider and Goldberg^26^ examined composite scales designed for preclinical AD and found that most of the newer composites did not include alternate forms to reduce practice effects. Only three of eleven reviewed scales contained partial alternate forms. Of note, the PACC,^3^ a battery currently being used in the A4 clinical trial and validated in multiple clinical trials, has limited alternate forms and was found to exhibit significant practice effects in the majority of study participants.^33^ The review paper also found that eight of the eleven composites reviewed include tests of orientation, despite findings that they are prone to substantial ceiling effects in individuals with preclinical AD.^34^ Surprisingly, ceiling effects, floor effects, and other psychometric properties like test-retest reliability were not reported in the majority of the studies reviewed. Additionally, nearly all recent composites have not been validated psychometrically as a composite; rather, many used single test psychometrics collected in many different “normative” groups over many years and have not demonstrated that the composite score outperforms individual test scores.

These findings indicate the need for composite measures with more robust psychometric properties, less redundancy, co-normed tests, and attenuated practice and ceiling effects. This study seeks to validate the composite scores derived from the NPE and CFAS, while also employing two well established scales, the PACC composite and the ADAS-Cog^4^, whose items assess multiple domains including memory, orientation, praxis, and naming.

We emphasize that while the NPE is designed to be a composite measure and that it is co-normed, its individual tests and empirically driven performance domains (executive and working memory function, memory, speed) can be additionally used to assess specific mechanisms based on disease (e.g., assessing episodic memory for AD pathology) and treatment.

### Strengths & Limitations

This is the first multicenter study designed as a clinical trial to validate a set of measures for use in AD clinical trials. The novel measures are validated within a clinical trials armature but also follow the innovative “intent-to-test” design in which participants are randomly assigned to a novel measures or established measures group that then undergoes serial assessment. Further, the multi-site design serves as a check on reproducibility and will provide data on site-related variance. A limitation of this study is that it does not include individuals in late-stage MCI given the focus on a pre-clinical population. The data set also does not include amyloid beta as one of its biomarkers, although the onset of amyloid positivity can be inferred from other measures we collect (e.g., ApoE genotype).

## Conclusions

Results from this study may support validation of two new composite measures, the NPE battery and the CFAS, while also yielding valuable data for the field. If validated, these measures may advance the clinical testing of pharmacological and non-pharmacological interventions for individuals across the spectrum of early stage (i.e., preclinical and MCI) AD. We also expect to see a shift toward remote administration of neuropsychological measures in a post-COVID-19 world. Considering that the majority of the novel tasks discussed here are delivered via laptop in clinic, these measures should be adaptable for remote administration.

## Data Availability

The study is still in recruitment. Data are not yet available.

## Authors’ Contributions

TEG designed the study and edited the manuscript. SAB and HRC wrote the first draft. HK, SL, AC, HA, AMR, KI, AMB, DPD, PDH, LSS and TEG scientifically edited the manuscript.

## Ethics Approval

This study is approved by the NYSPI IRB, the Feinstein Institute for Medical Research IRB, the University of Southern California Keck School of Medicine IRB, and the University of Miami Miller School of Medicine IRB.

## Data Sharing Statement

The study is still in recruitment. Data are not yet available.

